# Tuberculosis prevalence and incidence rates from repeated population-based screening in a district in Ethiopia: a prospective cohort study

**DOI:** 10.1101/2022.11.26.22282775

**Authors:** Abiot Bezabeh Banti, Sven Gudmund Hinderaker, Brita Askeland Winje, Einar Heldal, Markos Abebe, Mesay Hailu Dangisso, Daniel Gemechu Datiko

## Abstract

**Objective:** In Ethiopia, a third of the estimated tuberculosis cases are not detected or reported. Incidence estimates are inaccurate and are rarely measured directly. Any tuberculosis program will miss some tuberculosis cases and assessing the ‘‘real’’ incidence under program conditions is useful to understand the situation. This study aimed to measure the prevalence and incidence of pulmonary tuberculosis based on three rounds of household visits in the adult population of Dale in Ethiopia.

**Design:** A prospective population-based cohort study.

**Setting:** Every household in Dale district was visited three times at 4-month interval over 12 months.

**Participants:** Individuals aged 15 and more years were followed.

**Outcome measures:** Microscopy smear-positive PTB (PTB s+), bacteriologically confirmed PTB (PTB b+) by microscopy, GeneXpert, or culture, and clinically diagnosed PTB (PTB c+).

**Results:** Among 136,181 individuals, 2052 had presumptive TB in round1, including 93 with PTB s+, 98 with PTB b+, and 24 with PTB c+; adding those with PTB who were already on treatment, the total number of PTB was 201, and the prevalence was 147 (95%CI: 127–168)/100,000 population. Out of all detected PTB patients by screening, PTB s+ was 65%, PTB b+ was 67%, and PTB c+ was 44%. During 96,388 person-years follow-up after round1 to round3 end, 1909 had presumptive TB, 320 had PTB, and the total incidence of PTB was 332 (95%CI: 297–370)/100,000 person-years, while the incidence of PTB s+, PTB b+, and PTB c+ was 230 (95%CI: 201–262), 263 (95%CI: 232–297), and 68 (95%CI: 53–86)/100,000 person-years, respectively.

**Conclusion:** The prevalence of PTB s+ was similar to the national survey and local studies, but only a third of prevalent PTB cases had been notified. The incidence rate was highest in those aged 25–34 years, indicating ongoing transmission. Finding missing people with TB through repeated screening can reduce transmission.

**Strengths and limitations of this study:**

- This study included a large sample and worked closely with the national tuberculosis program and the community structures that contributed to its sustainability.
- We report not only tuberculosis prevalence but also incidence based on three rounds of household visits, which is often not reported elsewhere.
- The study screened a very high proportion of households in the district, giving an accurate measure of the burden of tuberculosis, where the probability of missing people with TB is small.
- Tuberculosis cases notified were chosen the year before the study because we could not separate the patients identified by the screening from those who attended the health services themselves.
- We did not perform GeneXpert and culture for all smear-positive tuberculosis cases, so there was a difference between microscopy smear-positive and bacteriologically confirmed tuberculosis cases, which was very small.

## BACKGROUND

Tuberculosis (TB) is an infectious disease transmitted by mycobacterial droplets from coughing patients. The basis of TB control, embodied in the global End TB strategy, is to reduce the sources of infection by curing infected individuals with standard treatment. Effective anti-TB drugs to cure TB have helped to reduce the incidence of TB worldwide. The estimated global incidence of TB has recently decreased; from 2015 to 2019, the incidence decreased by 9% (140 to 130 per 100,000 population), which is 11% less than the End TB targets set for 2020. The incidence of TB varies globally, with 86% of all estimated TB cases occurring in 30 countries. Africa has the highest estimated TB incidence rate, with 226 per 100,000 population in 2019. The TB incidence rate decline in Africa from 2015 to 2019 was 16%, which is higher than the average global decline but still not fast enough to reach the Sustainable Development Goal milestones. ^1-4^ Ethiopia achieved the first milestone of End TB with a 31% decrease in the estimated incidence rate from 2015 to 2020 (192 to 132 per 100,000 population).

The national TB survey in Ethiopia in 2010 reported *prevalence* estimates of 108 per 100,000 population for smear-positive pulmonary tuberculosis (PTB s+) and 277 per 100,000 population for bacteriologically confirmed pulmonary tuberculosis (PTB b+).^5^ Only two studies have investigated the *incidence*. A study conducted in a central district of Ethiopia in 2013 reported that the incidence of PTB s+ was 214 per 100,000 person-years and that of PTB b+ was 232 per 100,000 person-years,^6^ while a study in the northern part of the country in 2011 reported that the incidence of PTB s+ was 311 per 100,000 person-years.^7^ However, notifications in the country indicate that a third of TB cases are still undetected, and more than half of them are in those aged 15–34 years old. Hence, the true incidence of TB is unknown and under-researched because generating these estimates requires massive resources.^8 9^

Therefore, locally available data using the existing health system may help to understand the epidemiology of the disease as well as to validate national estimates, since these are not accurate in areas with disparities in health outcomes and program implementation. The National Tuberculosis and Leprosy Program (NTLP) aimed to improve TB case detection at the subnational level by the quarterly follow-up of contacts and individuals with symptoms via providing interventions under programmatic conditions using the current community program approach, but this has not been realized. Recent reports show decreases in the estimated incidence and reports of TB cases, possibly due to active case finding with health extension workers (HEWs).^3^ These are trained providers and part of primary health care services in Ethiopia, and they support treatment follow-up and patient adherence in the community for those enrolled in care. The “missed” or “diagnosed late” individuals are important sources of TB transmission.^5 10^ Repeated assessment of subnational population-level incidence and prevalence patterns may help the program quantify the impact of the control efforts to support local decision making.^2 4 5 10-13^

In the Dale district, there have been active TB case-finding activities, but their impact has not been sufficiently described and documented. Hence, this study aimed to measure the prevalence and incidence of pulmonary TB (PTB) through repeated population-based screening in the adult population of the Dale district in southern Ethiopia.

## METHODS

### Design and setting

This was a population-based prospective study with repeated screening of the population in three consecutive rounds of household visits in the Dale district of southern Ethiopia from October 2016 to September 2017. The Dale district is a densely populated rural community divided into 36 rural administrative units (*kebeles*). The study area has 10 health centers, two clinics, and 36 health posts; TB care follows the End TB strategy ^14^. Since 2010, the nongovernmental organization REACH Ethiopia has been developing an innovative model using close-to-community providers, including in the Dale district, contributing to community TB care implementation at the national level in Ethiopia.^6 15^

Aggregate notification data were obtained from the NTLP from 2011 to 2018. Quarterly TB data were used to track those notified with PTB s+ or *clinically diagnosed tuberculosis (PTB c+)* before, during, and after the study period to evaluate the impact of interventions on routine TB notification. Additionally, the number of patients already on TB treatment at the time of round 1 screening, and therefore were neither screened nor tested by the project, was determined. The project ensured that all TB patients identified through the project were entered into the TB register.^16^ We used the Strengthening the Reporting of Observational Studies in Epidemiology cohort reporting guidelines (File S1).^17^

### Patient and Public Involvement

No patients were involved in setting the research question or the outcomes, the conduct, interpretation, writing or dissemination of the results. The study was conducted in close collaboration with the woreda TB and leprosy programme in Dale. They were informed about the study in advance and were actively involved throughout the design and implementation phases and in dissemination of the results. The project team organized consultative meetings with participants from regional, zonal and woreda level organisations, non-governmental organizations, and religious institutions throughout the full study period. Village-women were included as representatives for the population in Dale.

### Data collection and participants

All residents aged 15 years or older were considered the source population. Those already on TB treatment at the time of the survey were not included in the screening data. However, aggregate surveillance data from the NTLP on the number of patients already on TB treatment were added to those detected by screening in the prevalence calculation to facilitate comparison with other studies. The total population denominator was 267,393, and the population aged 15 years or older was 136,181, based on official data from the Central Statistics Agency in Ethiopia (estimated from a population census of 2007, with an annual growth rate of 3%).^18^ The project team prepared the primary health care services for the study and created awareness about the study in the community. The NTLP at various levels was consulted in writing and meetings to ensure local ownership and compliance with local guidelines. A TB expert supervised the field activity throughout the study period, and specific training was given to HEWs, personnel responsible for TB control, and laboratory technicians to maintain the quality of project implementation in the field.

Pretested and semi-structured questionnaires were used to identify individuals with symptoms compatible with PTB and to collect clinical, demographic, and socioeconomic data (File S2). A persistent cough for 14 days or more with or without *hemoptysis, weight loss, fever, night sweats, chest pain, or difficulty breathing* was taken as a sign of presumptive PTB. Individuals identified as having presumptive TB were asked to come to the health post to provide sputum. The sputum samples were collected and transported to the health center for smear microscopy. Once identified by the study, participants received care, free of charge, as in routine management. If individuals were diagnosed with TB, they were treated according to the NTLP guidelines.^19^ For patients diagnosed with drug-resistant TB, second-line drugs and follow-up were available at Yirgalem Hospital. Individuals diagnosed with other diseases were referred for treatment. To ensure the quality of the laboratory findings, 50% of the slides were randomly selected from participating catchment facilities and retested at the regional laboratory. Smear-positive samples were further validated by GeneXpert.

### Operational definitions

The term “new presumptive” was used the first time an individual was identified in the study as having presumptive TB. Those with presumptive TB who tested negative for TB were included in the denominator for the consecutive rounds. If they were identified as having presumptive TB in a consecutive round of household visits (more than 30 days apart), they were classified as “previous presumptive with a new episode.” Individuals who were tested several times within the same round of household visits (4-month period) were classified as belonging to the same presumptive episode, as each household was only visited once in each round of household visits. PTB s+ was defined as at least one positive result by smear microscopy. PTB b+ included having at least one positive result by smear microscopy, GeneXpert, or culture. PTB c+ was based on a clinical decision to start anti-TB treatment in those with persistent symptoms and negative bacteriological results; this diagnosis was usually (if not always) also supported by radiological findings. PTB b+ and PTB c+ both indicated a diagnosis of PTB. The “screening prevalence” of PTB was calculated based on individuals identified with PTB in the first round (October 2016 to January 2017) of household visits divided by the total adult population (per 100,000 population). The “total prevalence” also included patients who were already on TB treatment at the time of the first screening round. The incidence was calculated based on the number of individuals newly identified with PTB in the second (February to May 2017) and third (June to September 2017) rounds of household visits divided by the person-years observation time (per 100,000 person-years). Person-years were calculated from the date of enrolment for screening to TB diagnosis or the end of follow-up on September 31, 2017, for all study participants. ‘‘Notification’’ (number of notified cases) is the aggregate data from the health facility’s TB register. Patients detected by screening could not be separated from those who attended health services themselves during the study. Therefore, the four quarters before the study were chosen as notification when comparing the prevalence and the incidence.

### Diagnosis

For diagnosis, two spot sputum samples were collected according to standard procedures. Sputum smear-positive samples were sent for confirmation of diagnosis by GeneXpert at Yirgalem Hospital and by culture at the Armauer Hansen Research Institute in Addis Ababa. In addition, smear-negative samples from selected individuals were sent for analysis by GeneXpert and cultured in line with NTPL guidelines. Data on TB diagnosis, treatment, and treatment outcome were obtained from the TB register in the health center. It was not possible to disentangle those identified through the household screening from those who approached the health center on their own initiative. Individuals with chest X-ray abnormalities^20^ were transferred to Yirgalem Hospital for clinical diagnosis of TB.

### Data quality and analysis

Data were entered into Excel (Microsoft® Office Proofing Tool 2016, Microsoft Corporation). The data quality was assessed by frequency distributions, cross-tabulations, and double-entry of a 10% random sample of the data set. The difference between the first and second data entries was 0.1%, and no systematic errors were detected. Stata version 14 and Openepi^21^ were used for analyses.

## RESULTS

### Population-based screening

In this study, coverage for screening was high across all three rounds of household visits, with 98%, 96%, and 97% coverage in rounds1, 2, and 3, respectively. A flow chart of the three rounds of household screening is presented in Figure 1. We screened 45,672 household with 136,181 adults and identified 3,746 individuals with presumptive PTB during the study period. Among them, 2052 were identified as having presumptive PTB in the initial round of household visits. The prevalence of presumptive PTB was 1507 per 100,000 population, and it increased with age. The number of individuals presumed to have PTB was the highest in the first round of household visits and halved in the two consecutive rounds (Table S1). The opposite was observed for PTB cases, which were higher in the second and third round. Seventeen TB cases with symptoms of PTB were eventually diagnosed with extrapulmonary TB but not PTB. These were excluded from the study (Table S1). The proportions of PTB cases among individuals with presumptive TB were 6%, 17%, and 16% across the three rounds; the proportions of individuals with PTB b+ were 5%, 12%, and 15%, respectively; and the proportions of PTB c+ were 5%, 10%, and 13%, respectively. The proportion of PTB among individuals with presumptive TB who completed a course of broad-spectrum antibiotics and returned for another test was higher in the second and third rounds (14%, 41%, and 31% for rounds 1, 2, and 3, respectively). Out of 994 individuals with presumptive TB identified in the third round, 351 had the results of all three tests, with 21 (6.0%) smear-positive by microscopy, 26 (7.4%) positive by GeneXpert, and 27 (7.7%) positive by culture. Four individuals were identified with rifampicin resistance among 263 PTB b+ cases with rifampicin resistance test results. Females had a significantly higher prevalence and incidence of presumptive TB, a lower proportion with PTB b+ cases, and a lower prevalence and incidence of PTB than males (but not significant) (Table S2, S3 and S4).

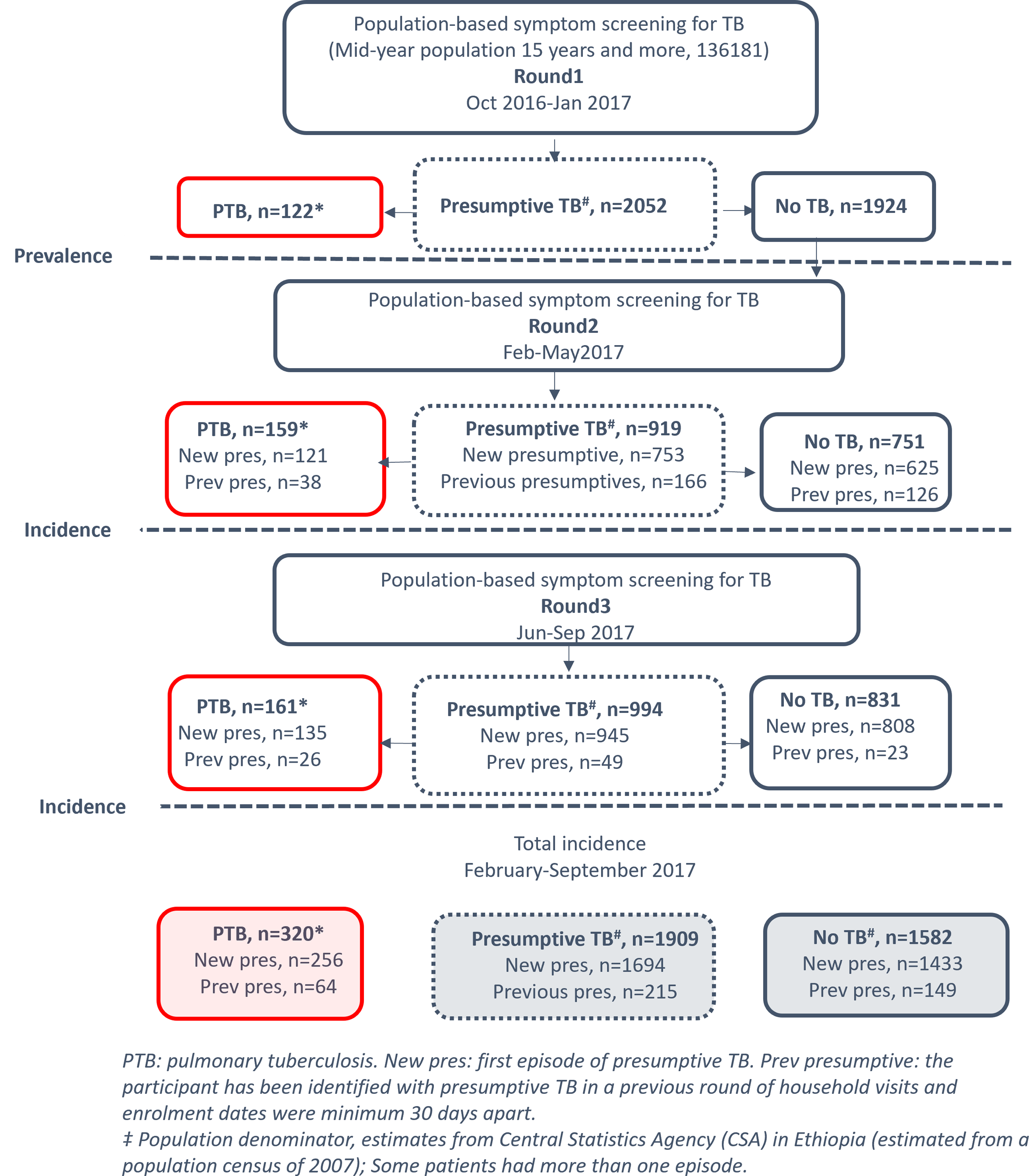

### Prevalence of PTB

Table 2 shows the total prevalence of PTB in the study area in round 1 household visits. Among the population of 136,181, a total of 2052 had presumptive TB, 93 were diagnosed with PTB s+, 98 with PTB b+, and 24 with PTB c+. The total number of PTB patients detected by screening was 122; adding 49 PTB b+ (all of them smear-positive) and 30 PTB c+ patients who were already on TB treatment at the first round, the total number of PTB cases was 201, and the prevalence rate of PTB was 147 (95%CI: 127–168) per 100,000 population. In those detected by screening, the prevalence rates of PTB b+ and PTB c+ were 72 (95%CI: 57– 86) and 17 (95% CI: 10–24) per 100,000 population, respectively. The prevalence rate of PTB s+ was 68 (95%CI: 54–82) per 100,000 population from screening and 104 (95% CI: 87–121) per 100,000 population if patients already on treatment were included. The percentages of patients detected by screening were as follows: PTB s+, 65%; PTB b+, 67%; and PTB c+, 44%. The proportion of PTB detected by screening ranged from 38% in Mesenkala to 91% in Kege, but there was no statistically significant distinction between the areas (Supplementary file S4). Among both PTB b+ and PTB c+ patients detected, only one out of three had been identified by the health system (Table S5).

**Table 1.** Results of three rounds of testing individuals with presumptive tuberculosis in Dale, 2016–2017.

| Household visits |  | Presumptive TB episodes<sup>#</sup>, *n* | PTB, *n* | PTB b+, *n* | PTB c+, *n* | % presumptive TB with TB |  |  |
| --- | --- | --- | --- | --- | --- | --- | --- | --- |
|  |  |  |  |  |  | PTB<sup>1</sup> | PTB b+<sup>1</sup> | PTB c+<sup>1</sup> |
| Round 1 | New presumptive; only tested once | 1729 | 76 | 71 | 5 | 4% | 4% | 0.3% |
|  | New presumptive; TB with a repeat test | 323 | 46 | 27 | 19 | 14% | 8% | 5.9% |
|  | New presumptive; all* | 2052 | 122 | 98 | 24 | 6% | 5% | 1.2% |
| Round 2 | New presumptive; only tested once | 630 | 72 | 71 | 1 | 11% | 11% | 0.2% |
|  | New presumptive; TB with a repeat test | 119 | 49 | 19 | 30 | 41% | 16% | 25.2% |
|  | New presumptive; all* | 753 | 121 | 90 | 31 | 16% | 12% | 4.1% |
|  | Previous presumptive | 166 | 38 | 19 | 19 | 23% | 11% | 11.4% |
|  | Total | 919 | 159 | 109 | 50 | 17% | 12% | 5.4% |
| Round 3 | New presumptive; only tested once | 900 | 121 | 119 | 2 | 13% | 13% | 0.2% |
|  | New presumptive; TB with a repeat test | 45 | 14 | 11 | 3 | 31% | 24% | 6.7% |
|  | New presumptive; all* | 945 | 135 | 130 | 5 | 14% | 14% | 0.5% |
|  | Previous presumptive | 49 | 26 | 15 | 11 | 53% | 31% | 22.4% |
|  | Total | 994 | 161 | 145 | 16 | 16% | 15% | 1.6% |
| Rounds 1+2+3 | New presumptive | 3746 | 378 | 318 | 60 | 10% | 8% | 2.0% |
|  | Previous presumptive | 215 | 64 | 34 | 30 | 30% | 16% | 14.0% |

\*, New presumptive all means only tested once plus repeat tests within the same round. TB, tuberculosis; PTB, pulmonary TB, PTB b+, bacteriologically confirmed PTB; PTB c+, clinically diagnosed PTB; 1, PTB means the proportion of presumptive TB cases with a positive bacteriological test result plus clinically diagnosed PTB. Individuals with pulmonary presumptive TB who were eventually diagnosed with extrapulmonary TB, but without PTB. #, the number of episodes is higher than the number of individuals as some were identified more than once.

**Table 2.** Prevalence of individuals with presumptive tuberculosis (TB), total prevalence, and proportion detected with pulmonary TB in the first round of visits in Dale, October 2016–January 2017.

| Covariate | Population | Presumptive TB |  | Number of PTB during screening |  | Number of PTB before screening |  | Total prevalence of PTB |  |  |  | Total detected | % detected by screening |  |  |
| --- | --- | --- | --- | --- | --- | --- | --- | --- | --- | --- | --- | --- | --- | --- | --- |
|  |  | *n* | per 100,000 | PTB b+ | PTB c+ | PTB b+ | PTB c+ | PTB b+ | per 100,000 | PTB c+ | per 100,000 |  | PTB b+ | PTB c+ | Total |
| Total | 136,181 | 2052 | 1507 (1442–1572) | 98 | 24 | 49 | 30 | 147 | 108 (90–125) | 54 | 39 (29–50) | 201 | 67% | 44% | 61% |
| Age in years |  |  |  |  |  |  |  |  |  |  |  |  |  |  |  |
| 15–24 | 49,735 | 294 | 591 (527–661) | 27 | 7 | 23 | 5 | 50 | 100 (76–132) | 12 | 24 (13–41) | 62 | 54% | 58% | 55% |
| 25–34 | 33,958 | 395 | 1163 (1053–1281) | 31 | 7 | 15 | 6 | 46 | 135 (100–179) | 13 | 38 (21–64) | 59 | 67% | 54% | 64% |
| 35–44 | 23,797 | 437 | 1836 (1672–2013) | 26 | 2 | 4 | 2 | 30 | 126 (86–177) | 4 | 17 (5–40) | 34 | 87% | 50% | 82% |
| 45–54 | 13,369 | 432 | 3231 (2941–3541) | 7 | 4 | 4 | 5 | 11 | 82 (43–143) | 9 | 67 (33–123) | 20 | 64% | 44% | 55% |
| 55+ | 15,319 | 494 | 3225 (2954–3514) | 7 | 4 | 3 | 12 | 10 | 65 (33–116) | 16 | 104 (61–166) | 26 | 70% | 25% | 42% |
| Sex |  |  |  |  |  |  |  |  |  |  |  |  |  |  |  |
| Male | 67,681 | 847 | 1251 (1170–1337) | 56 | 15 | 27 | 17 | 83 | 122 (98–151) | 32 | 47 (33–66) | 115 | 67% | 47% | 62% |
| Female | 68,500 | 1205 | 1759 (1662–1860) | 42 | 9 | 22 | 13 | 64 | 93 (72–118) | 22 | 32 (21–48) | 86 | 66% | 41% | 59% |

TB, tuberculosis; PTB, pulmonary TB; PTB b+, bacteriologically confirmed PTB; PTB c+, clinically diagnosed PTB.

### Incidence of PTB

Table 3 shows the PTB incidence in the study area. The overall follow-up period (round 2 plus round 3) included an observation time of 96,388 person-years, with 1,909 individuals identified as having presumptive TB, 254 having PTB b+ (including 222 smear-positive), and 66 having PTB c+. The total number of PTB cases was 320. The incidence of PTB was 332 (95% CI: 297–370) per 100,000 person-years; specifically, the incidence rates of PTB s+, PTB b+, and PTB c+ were 230 (95% CI: 201–262), 263 (95% CI: 232–297), and 68 (95% CI: 53–86) per 100,000 person-years, respectively. Importantly, the incidence rates of PTB s+ and PTB b+ were the highest in those aged 25–34 years old (Table S6). In addition, PTB rate in previous presumptive episode group, followed for 2,997 person-years, was 2,135 per 100,000 person-years.

**Table 3.**
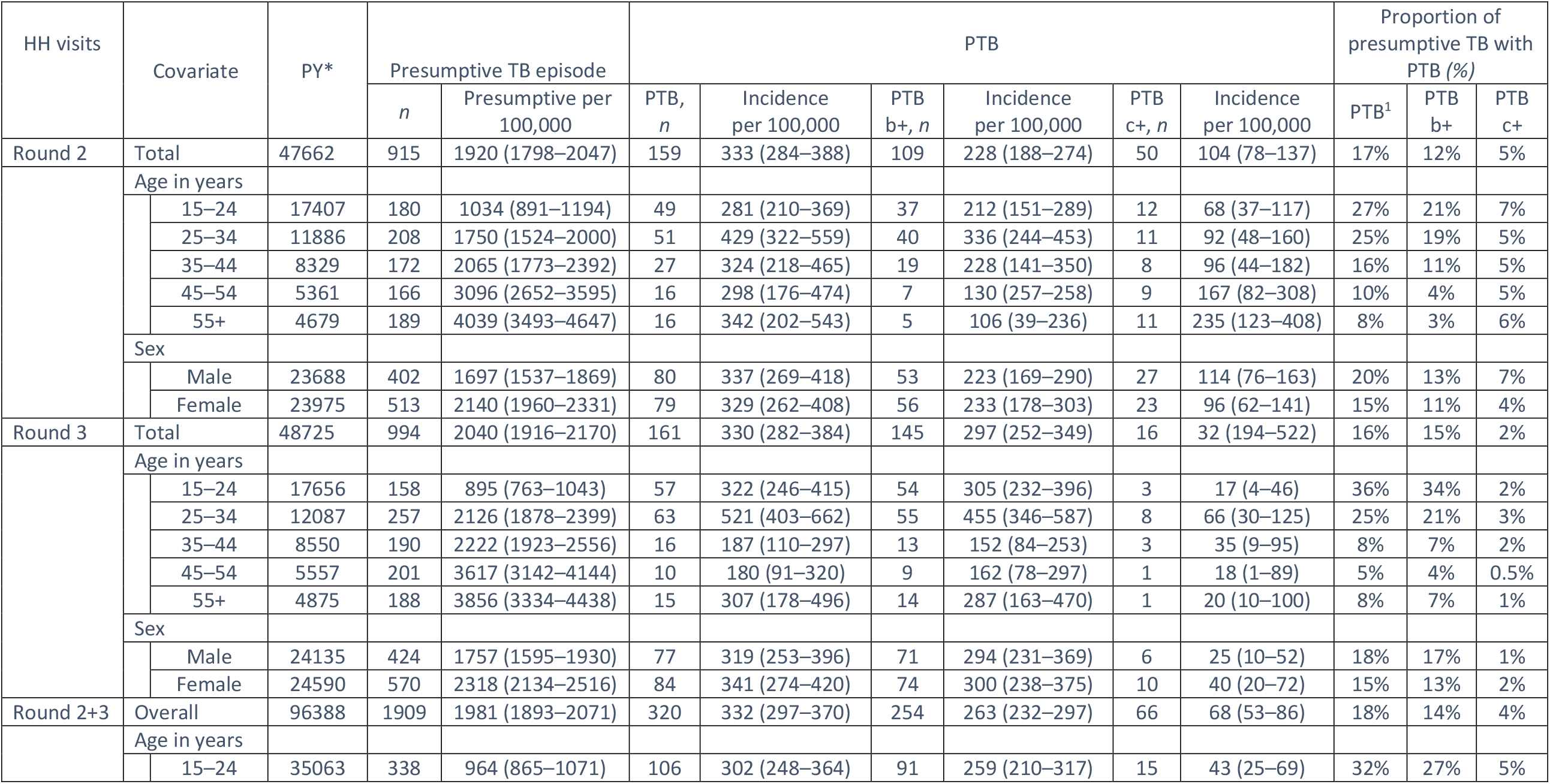
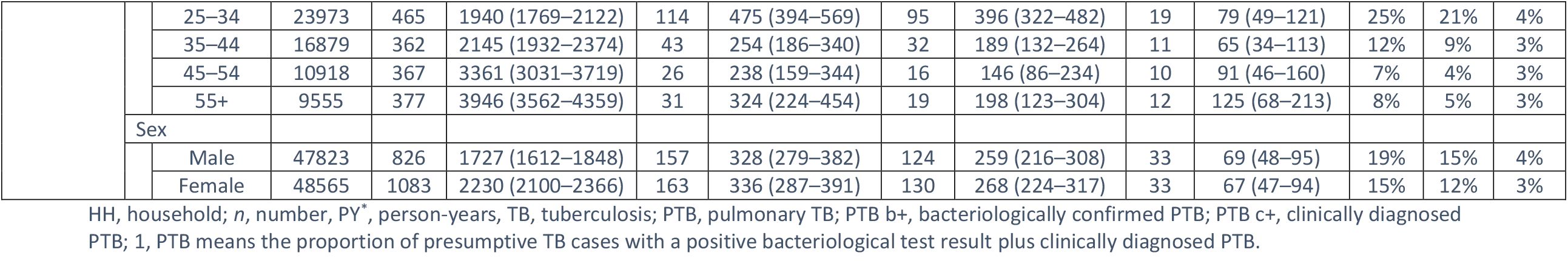
Incidence of presumptive and pulmonary tuberculosis in the adult population in Dale, February–September 2017.

### PTB rate ratio comparison by sex

Table 4 shows that the ratios of the notification rate to the total prevalence rate (including cases already on TB treatment in round 1 of the survey) as well as to the incidence rates for PTB b+, PTB c+, and PTB s+. The prevalence-to-notification rate ratios for PTB s+ were 1.28:1 for all adults, 1.2:1 for males, and 1.4:1 for females. The ratios of the prevalence rate to the incidence rate were 0.45:1 for all adults, 0.52:1 for males, and 0.40:1 for females. The ratios of the notification rate to the incidence rate of PTB b+ were 0.35:1 for all adults, 0.42:1 for males, and 0.28:1 for females.

**Table 4.** Measurements of bacteriologically and clinically diagnosed pulmonary tuberculosis, by sex, in Dale.

| Indicator | Classification | PTB b+ per 100,000<br>(95% CI) | PTB c+ per 100,000<br>(95% CI) | PTB s+ per 100,000<br>(95% CI) |
| --- | --- | --- | --- | --- |
| Rates |  |  |  |  |
| Notification rate before the study (4<sup>th</sup> quarter 2015 to 3<sup>rd</sup> quarter 2016) | All | 81 (66–96) | 52 (40–64) | 81 (66–96) |
|  | Male | 96 (75–122) | 65 (48–87) | 96 (75–122) |
|  | Female | 66 (48–87) | 32 (20–48) | 66 (48–87) |
| Total prevalence rate | All | 108 (90–125) | 39 (29–50) | 104 (87–121) |
|  | Male | 122 (98–151) | 47 (33–66) | 117 (93–144) |
|  | Female | 93 (72–118) | 32 (21–48) | 92 (71–117) |
| Incidence rate | All | 263 (232–297) | 68 (53–86) | 230 (201–262) |
|  | Male | 259 (216–307) | 69 (48–95) | 226 (187–271) |
|  | Female | 268 (224–317) | 67 (47–94) | 235 (195–281) |
| Rate ratios |  |  |  |  |
| Total prevalence-to-notification ratio | All | 1.3 (1.09–1.6) | 0.75 (0.54–1.01) | 1.28 (1.05–1.55) |
|  | Male | 1.27 (1.06–1.5) | 0.72 (0.60–0.82) | 1.2 (1.0–1.4) |
|  | Female | 1.4 (1.14–1.7) | 1.0 (0.91–1.0) | 1.4 (1.1–1.7) |
| Total prevalence-to-incidence ratio | All | 0.41 (0.33–0.49) | 0.57 (0.45–0.68) | 0.45 (0.37–0.54) |
|  | Male | 0.47 (0.41–0.53) | 0.68 (0.56–0.78) | 0.52 (0.43–0.62) |
|  | Female | 0.35 (0.29–0.40) | 0.48 (0.36–0.59) | 0.4 (0.32–0.48) |
| Notification-to-incidence ratio | All | 0.31 (0.25–0.36) | 0.76 (0.65–0.85) | 0.35 (0.28–0.43) |
|  | Male | 0.37 (0.31–0.43) | 0.94 (0.86–0.98) | 0.42 (0.35–0.52) |
|  | Female | 0.24 (0.2–0.30) | 0.48 (0.36–0.59) | 0.28 (0.22–0.35) |

PTB b+, bacteriologically diagnosed pulmonary tuberculosis; PTB c+, clinically diagnosed pulmonary tuberculosis; PTB s+, smear-positive pulmonary tuberculosis; CI, confidence interval.

### PTB notification trend by year

Figure 2a shows that the initial high level of notification declined during the previous project of screening and that it increased moderately during the current project. In addition, Figure 2b shows the notification rates in the year (four quarters) before, during, and following the intervention. The PTB s+ notification rates were 81 (95%CI: 66–96), 231 (95%CI: 206–257), and 150 (95% CI: 130–170) per 100,000 population, respectively. The notification rates increased threefold during the project year and then decreased after the project but were higher than before the project year started. The notification rates had wide ranges between catchment areas, particularly before the start of the current project [Table S7, S8 and S9].

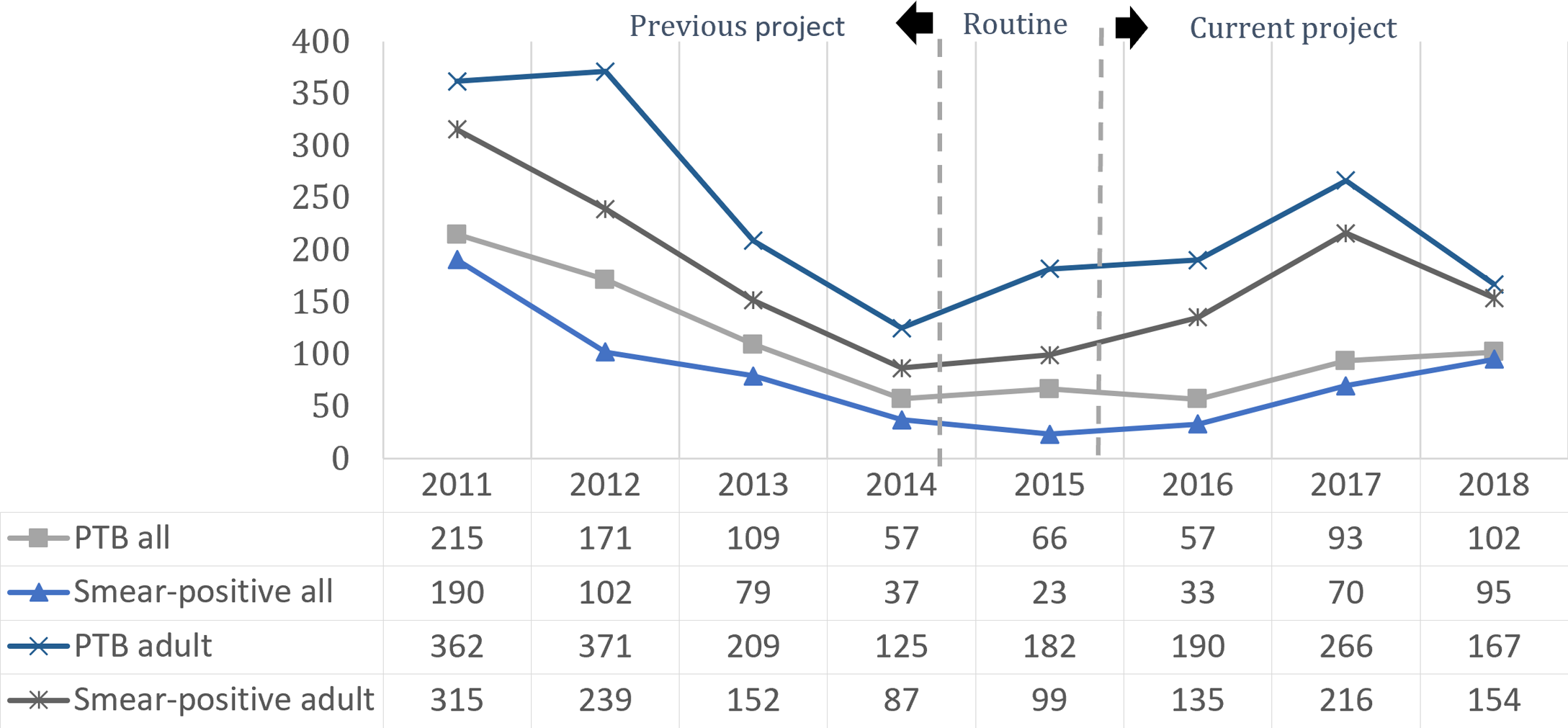
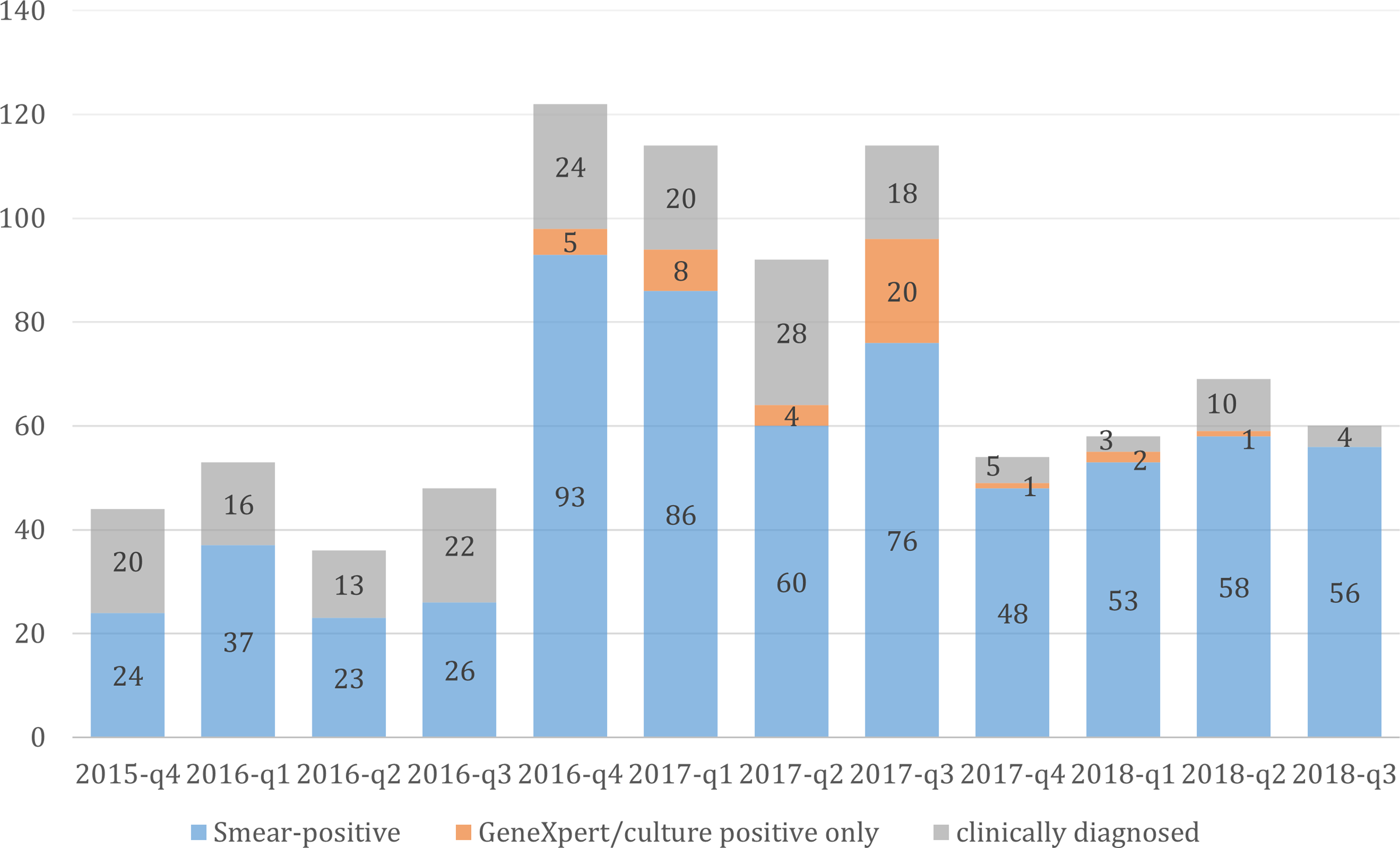

## DISCUSSION

This systematic population-based TB screening in a district of southern Ethiopia identified a point prevalence of smear-positive TB of 104 per 100,000 population, similar to that found by the 2010 national prevalence survey in Ethiopia (108 per 100,000 population).^5^ Only a third of individuals with TB were detected by the routine NTLP services. The incidence of PTB was 332 per 100,000 person-years; out of them, 263 were PTB b+ and 230 were PTB s+. The incidence of PTB s+ and PTB b+ was the highest among those with persistent symptoms and in those aged 25–34 years old, suggesting continued TB transmission in the community.

Our study showed a prevalence similar to that found in a 2013 local survey from central Ethiopia, with a PTB s+ prevalence rate of 109 (67–150) per 100,000 population,^6^ while other studies have reported a higher prevalence of PTB s+: 169 per 100,000 population in northern Ethiopia in 2011^22^ and 139 per 100,000 population in southern Ethiopia in 2016.^23^ Other studies from different rural areas in Ethiopia have found PTB s+ prevalence rates of 78–174 per 100,000 population, demonstrating how the extent of the disease varies across the country.^13 24-26^ Compared to our data from Ethiopia, a systematic review of national TB prevalence surveys in Africa has revealed a higher prevalence of PTB s+ in Kenya (230 per 100,000) and Uganda (174 per 100,000), but a comparable prevalence in Ghana (111 per 100,000), and a lower prevalence in Gambia (90 per 100,000) and Rwanda (74 per 100,000).^27^ The difference in prevalence rates across studies reflects variation in methods, year of study, and location; for example, the national survey in Ethiopia used chest x-rays for screening in addition to smear microscopy and culture, while others, including the current study, used symptom screening. Studies based on smear microscopy alone have lower prevalence rates than studies using the more sensitive techniques of GeneXpert and culture.^22 23 28^

In this study, two out of three individuals with PTB s+ remained undetected in the communities, which is in agreement with a systematic review showing a point prevalence of undiagnosed PTB s+ of 79 (56–113) per 100,000 population, with an active-to-passive case finding ratio of 2.3:1,^28^ which is also comparable to our prevalence of 72 (57–86) per 100,000 population. Besides, two individuals out of five with PTB c+ remained undetected. PTB c+ contributed 20–36% to the total prevalence and incidence of PTB in our study; 44% of PTB c+ cases were detected by screening. In general, all PTB c+ diagnoses were by chest x-ray, but the overall incidence of PTB was not too high; this can be a cause for concern, and physicians may have diagnosed PTB c+ in more severely ill older people than in people with fewer symptoms. Overall, project-based activities performed in the study setting with active TB case finding increase TB detection through community engagement with HEWs, thereby improving community awareness and access to TB treatment.^29 30^ A lower prevalence of HIV and co-infections in the region^31^ may have contributed to a lower prevalence compared to the other studies.

More women were found to be symptomatic during round 1, and the prevalence of presumptive TB was significantly greater among women than men. However, the prevalence of TB was not statistically different by sex in this study. Women accounted for only 44% of the nationally notified cases in 2020 in Ethiopia, and such trends persisted for several years. TB mainly affects women due to socioeconomic disadvantages and domestic responsibilities.^11^ The catchment areas of Gidamo, Kege, and Boa had a higher prevalence of presumptive TB than the other catchment areas, but Gidamo had the highest prevalence of presumptive TB in the first round: 2166 (1853–2516) per 100,000 population. However, a significant difference in the incidence across the study areas was not found, despite significant variation with wide ranges. Analysis at a lower level in the community may facilitate an understanding of different determinants of TB (i.e., barriers to TB services). The small numbers of cases in the subanalyses caused wide confidence intervals; therefore, the data must be interpretated with caution (Table S4).

The incidence of PTB s+ was 230 (201–262) per 100,000 person-years, which is similar to the most recent report, 214 (163–263) per 100,000 person-years for PTB s+ from central Ethiopia in 2013, but lower than 311 (240–382) per 100,000 person-years in northern Ethiopia in 2011;^6 7^ however, the confidence intervals overlap. The group of those aged 25–34 years old had the highest incidence of PTB s+ cases of any age group, but the incidence of PTB c+ showed no significant difference with age or sex; this may indicate heightened recent community transmission in this age group. The group of those aged 15–24 years old had a lower incidence [259 (210–317) per 100,000 person-years] than those aged 25–34 years old [396 (322– 482) per 100,000 person-years].

Individuals who had presumptive TB for more than one round of screening (previous presumptive) were more likely to have PTB b+ (16%) than a new presumptive TB case identified for the first time (8%).^32^ Similar trends were seen in a study in Guinea-Bissau, where smear-negative chronic coughers had a 5% higher smear positivity rate after a month than new presumptive TB cases.^33^ As expected, this study found more cases of PTB with a longer follow-up of chronic coughers after negative results than those initially identified with presumptive TB, thus emphasizing the need to reach this population with feasible interventions and follow-up that can improve the identification of more TB cases.^33 34^

Our study allowed us to directly calculate the prevalence-to-incidence ratio. In an ideal world, where all patients are detected early, reported quickly, and cured after six months of treatment, this ratio should be near 0.5. In our study of those with PTB s+, the ratios were 0.45:1 overall, 0.52:1 in males, and 0.4:1 in females; these findings are likely due to the systematic and repeated screening with high coverage. Our prevalence-to-incidence ratio was lower than that found in the only study reporting both prevalence and incidence rates in Ethiopia, which had a prevalence-to-incidence ratio of 0.6:1.^6^

In prevalence surveys, the true incidence is normally unknown, and notification rates are used to estimate the incidence while taking into consideration underdiagnoses and underreporting. The prevalence-to-notification rate ratio in PTB s+ cases in our study was 1.28:1, consistent with the finding that only one-third of TB patients were detected and notified; therefore, they had TB for a long time before diagnosis. In national surveys, the prevalence-to-notification rate ratios in PTB s+ patients have been reported to be 1.19:1 in Ethiopia, 0.62:1 in Gambia, and 5.8:1 in Nigeria.^27^

The notification rate increased threefold during the intervention year. Even if it decreased after the intervention, the notification rate remained at a high level most likely due to increased awareness about TB in the community. This underscores the value of repeated and continuous TB screening in the community to reduce ongoing transmission.

This study had two main strengths. First, the project worked in close collaboration with the NTLP, and very few people declined to participate. Due to the large study population, the data provide precise estimates of the prevalence and incidence of PTB. Second, the engagement of existing health care workers, health centers, district supervisors, and community structures increased the community involvement, thus contributing to its sustainability. There were also some challenges. Symptom-based TB screening was used, which identifies perhaps only half of the confirmed TB patients.^35^ In addition, since the patients detected by screening could not be separated from those who attended health services by themselves, the year before the study was chosen as “notification.” Furthermore, GeneXpert and culture were not performed for all PTB s+ cases, so the difference between PTB s+ and PTB b+ cases was very small.

The public health implication of this study is that two-thirds of symptomatic individuals with PTB were undetected. TB screening is important to understand who and where the missing symptomatic patients are as well as to improve access to and the functioning of health services, e.g., decentralized GeneXpert diagnostics or an improved sample transport system. Screening can only be performed periodically, since it is costly and time-consuming, and TB is constantly developing.

## CONCLUSIONS

The prevalence rate of PTB s+ was similar to those found in the national survey in 2010 as well as other local studies. Moreover, only a third of PTB cases were notified. The incidence rate of PTB was also similar to that most recently reported in 2013, and the incidence was the highest in those aged 25–34 years old. These findings indicate the ongoing transmission of PTB. Finding “missing” people with TB by using cost-effective interventions under programmatic conditions by engaging existing health system structures can reduce the “TB gaps,” decrease transmission, and contribute to a sustained decline in the incidence of TB in a society.

## Data Availability

The datasets generated and/or analysed during the current study are not publicly available to protect the confidentiality of the study subjects but are available from the corresponding author on reasonable request. To receive access to the data the applicant will need to provide an ethical approval from their IRB or equivalent body, and approval from the AHRI-ALERT and National Research Ethics Committees in Ethiopia.

## Abbreviations

HEWs: Health Extension Workers
NTLP: National Tuberculosis and Leprosy Program
PTB: pulmonary tuberculosis
PTB b+: bacteriologically confirmed pulmonary tuberculosis
PTB c+: clinically diagnosed pulmonary tuberculosis
PTB s+: smear-positive pulmonary tuberculosis
TB: tuberculosis

## Ethics approval and consent to participate

Ethics approval was obtained from the Armauer Hansen Research Institute-Alert Ethics Review Committee (PO12/15), the National Research Ethics Committee, Ethiopia (no 104/2016), and the Regional Committees for Medical and Health Research Ethics in Norway (2015/1006). The project received technical support from the Sidama Regional Health Bureau, formerly the Sidama Zone Health Department, the Dale District Health office, and from the health workers throughout the project period. Informed consent was also obtained at the time of interview and referral. A person who did not consent to participate and was already on TB treatment at the time of the survey was not eligible for screening activities. Informed consent was obtained from participants aged 15-17 years in addition to parental consent in accordance with Ethiopian guidelines.The project supported those individuals who could not afford the medical and transport costs during further investigation.

## Consent for publication

Not applicable

## Author contributions

BAW, DG, and EH wrote the protocol, AB, MA, and DG implemented the study in the community and collected the data. AB, DG, BAW, and EH supervised the fieldwork. AB, DG, BAW, and EH analyzed the data and drafted the manuscript. AB, DG, SGH, EH, MH, and BAW finalized the write-up. All authors reviewed and approved the final version of the manuscript.

## Acknowledgements

We are grateful to the University of Bergen for supporting the study. We thank Mr. Garedew Yeshitila for his continuous and close monitoring of the project in the field. We also would like to express our gratitude to the Sidama Regional Health Bureau, Yirgalem Hospital, Armauer Hansen Research Institute, and community members for their collaboration and support. We acknowledge the health professionals who worked closely with the researchers throughout the implementation of the project. We also thank the Norwegian Institute of Public Health and the Norwegian Health Association for funding this study.

## Supplementary appendix

- File S1 Strobe checklist.
- File S2 Adult symptom screening questionnaire.
- Table S1 Description of the screening flow with tuberculosis (TB) patients identified among those with presumptive TB in Dale from 2016 to 2017.
- Table S2 Screening prevalence of tuberculosis in round 1, by catchment area, in Dale from October 2016 to January 2017.
- Table S3 Proportion of pulmonary tuberculosis (TB) cases detected by screening and already on TB treatment at screening by *Kebele* in Dale from 2016 to 2017.
- Table S4 Proportion of pulmonary tuberculosis (TB) cases detected by screening and already on TB treatment at screening, by age and sex, in Dale from 2016 to 2017.
- Table S5 Screening and overall prevalence of pulmonary tuberculosis (including PTB s+) in round 1, by age and sex, in Dale from 2016 to 2017.
- Table S6 Incidence and proportion of smear-positive tuberculosis in the adult population in rounds 2 and 3, by age and sex, in Dale from February to September 2017.
- Table S7 Case notification per 100,000 population of bacteriologically diagnosed pulmonary tuberculosis in Dale from 2016 to 2018.
- Table S8 Case notification per 100,000 population of clinically diagnosed pulmonary tuberculosis in Dale from 2016 to 2018.
- Table S9. Case notification per 100,000 population of smear-positive pulmonary tuberculosis in Dale from 2016 to 2018.

## References

1. Riley RL MC, O’Grady F, Sultan LU, Wittstadt F, Shivpuri DN. Infectiousness of air from a tuberculosis ward; Ultraviolet irradiation of infected air comparative infectiousness of different patients,. Am Rev Respir Dis 1962; 1962;85: 511–25.

2. Rieder HL. Epidemiologic basis of tuberculosis control. Paris, Farance: International Union Against Tuberculosis and Lung Disease, 1999.

3. World Health Organization. Global Tuberculosis Report,, 2020.

4. World Health Organization. The end TB Strategy: Global strategy and targets for tuberculosis prevention, care and control after 2015 Geneva: Switherland; 2015 [

5. Kebede AH, Alebachew Z, Tsegaye F, et al. The first population-based national tuberculosis prevalence survey in Ethiopia, 2010-2011. IntJTubercLung Dis 2014;18(6):635–39. doi: 10.5588/ijtld.13.0417 [doi]

6. Hamusse S, Demissie M, Teshome D, et al. Prevalence and Incidence of Smear-Positive Pulmonary Tuberculosis in the Hetosa District of Arsi Zone, Oromia Regional State of Central Ethiopia. BMC infectious diseases 2017;17(1):214. doi: 10.1186/s12879-017-2321-0 [published Online First: 2017/03/18]

7. Tadesse T, Demissie M, Berhane Y, et al. Incidence of smear-positive tuberculosis in Dabat, northern Ethiopia. The international journal of tuberculosis and lung disease 2013;17(5):630–5. doi: 10.5588/ijtld.12.0449 [published Online First: 2013/04/12]

8. Dye C, Bassili A, Bierrenbach AL, et al. Measuring tuberculosis burden, trends, and the impact of control programmes. The Lancet Infectious diseases 2008;8(4):233–43. doi: 10.1016/s1473-3099(07)70291-8 [published Online First: 2008/01/19]

9. World Health Organization. Stop TB policy paper: TB impact measurement: policy and recommendations for how to assess the epidemiological burden of TB and the impact of TB control: World Health Organization 2009.

10. Federal Democratic Republic of Ethiopia Ministry of Health. National Strategic Plan Tuberculosis and Leprosy Control 2021-2026. Addis Ababa, Ethiopia;, 2021.

11. Federal Democratic Republic of Ethiopia Ministry of Health. National TB program report, 2020/21.

12. World Health Organization. Global tuberculosis report Geneva 2019 [Available from: https://apps.who.int/iris/bitstream/handle/10665/329368/9789241565714-eng.pdf?ua=12020.

13. Tadesse T, Demissie M, Berhane Y, et al. Two-thirds of smear-positive tuberculosis cases in the community were undiagnosed in Northwest Ethiopia: population based cross-sectional study. PLoSOne 2011;6(12):e28258. doi: 10.1371/journal.pone.0028258 [doi];PONE-D-11-19575 [pii]

14. World Health Organization. Implementing the end TB strategy: the essentials: World Health Organization, 2015.

15. Yassin MA, Datiko DG, Tulloch O, et al. Innovative community-based approaches doubled tuberculosis case notification and improve treatment outcome in Southern Ethiopia. 2013;8(5):e63174.

16. Dale Woreda Health Office. Tuberculosis and Leprosy Annual report from 2011-2018;, 2019.

17. von Elm E, Altman DG, Egger M, et al. The Strengthening the Reporting of Observational Studies in Epidemiology (STROBE) Statement: Guidelines for reporting observational studies. International Journal of Surgery 2014;12(12):1495–99. doi: https://doi.org/10.1016/j.ijsu.2014.07.013

18. Federal Democratic Republic of Ethiopia Central Statistics Agency. Summary and Statistical Report of the 2007 Population and Housing Census 2007 [Available from: https://www.ethiopianreview.com/pdf/001/Cen2007_firstdraft(1).pdf2021.

19. Federal Democratic Republic of Ethiopia Ministry of Health. Tuberculosis, leprosy and TB/HIV prevention and control program manual. Fifth edition ed: Ministry of Health 2012.

20. UNION. Desk-guide for diagnosis and management of TB in children. In: Diseases IUATaL, ed. Paris, France, 2010.

21. Dean AG SK, Soe MM. OpenEpi: Open Source Epidemiologic Statistics for Public Health, [updated updated 2013/04/06,. Version. https://www.OpenEpi.com, :[accessed 2019/07/24.

22. Berhe G, Enqueselassie F, Hailu E, et al. Population-based prevalence survey of tuberculosis in the Tigray region of Ethiopia. BMC infectious diseases 2013;13:448–48. doi: 10.1186/1471-2334-13-448

23. Merid Y, Mulate YW, Hailu M, et al. Population-based screening for pulmonary tuberculosis utilizing community health workers in Ethiopia. International journal of infectious diseases : IJID : official publication of the International Society for Infectious Diseases 2019;89:122–27. doi: 10.1016/j.ijid.2019.10.012 [published Online First: 2019/10/23]

24. Shargie EB, Yassin MA, Lindtjørn B. Prevalence of smear-positive pulmonary tuberculosis in a rural district of Ethiopia. The international journal of tuberculosis and lung disease : the official journal of the International Union against Tuberculosis and Lung Disease 2006;10(1):87–92. [published Online First: 2006/02/10]

25. Yimer S, Holm-Hansen C, Yimaldu T, et al. Evaluating an active case-finding strategy to identify smear-positive tuberculosis in rural Ethiopia. The international journal of tuberculosis and lung disease : the official journal of the International Union against Tuberculosis and Lung Disease 2009;13(11):1399–404. [published Online First: 2009/10/29]

26. Datiko DG, Guracha EA, Michael E, et al. Sub-national prevalence survey of tuberculosis in rural communities of Ethiopia. BMC public health 2019;19(1):295. doi: 10.1186/s12889-019-6620-9 [published Online First: 2019/03/15]

27. Law I, Floyd K, Group ATPS, et al. National tuberculosis prevalence surveys in Africa, 2008–2016: an overview of results and lessons learned. Tropical medicine & international health : TM & IH 2020;25(11):1308–27.

28. Arega B, Tilahun K, Minda A, et al. Prevalence rate of undiagnosed tuberculosis in the community in Ethiopia from 2001 to 2014: systematic review and meta-analysis. Archives of Public Health 2019;77(1):33. doi: 10.1186/s13690-019-0360-2

29. Datiko DG YM, Theobald SJ, et al. Health extesnsion workers improve tuberculosis case finding and treatment outcome in Ethiopia: a large scale implementation study. BMJ Global Health 2017

30. Tulloch O, Theobald S, Morishita F, et al. Patient and community experiences of tuberculosis diagnosis and care within a community-based intervention in Ethiopia: a qualitative study. BMC public health 2015;15:187. doi: 10.1186/s12889-015-1523-x [published Online First: 2015/04/18]

31. CSA and ICF. Ethiopian Demograpphic Health Survey 2016: HIV Report:. Addis Ababa, Ethiopia, and Rockville, Maryland, USA,: CSA and ICF, 2016.

32. Banti AB, Datiko DG, Hinderaker SG, et al. How many of persistent coughers have pulmonary tuberculosis? Population-based cohort study in Ethiopia. 2022;12(5):e058466. doi: 10.1136/bmjopen-2021-058466 %J BMJ Open

33. Porskrog A, Bjerregaard-Andersen M, Oliveira I, et al. Enhanced tuberculosis identification through 1-month follow-up of smear-negative tuberculosis suspects. The international journal of tuberculosis and lung disease : the official journal of the International Union against Tuberculosis and Lung Disease 2011;15(4):459–64. doi: 10.5588/ijtld.10.0353 [published Online First: 2011/03/15]

34. Keflie TS, Ameni G. Microscopic examination and smear negative pulmonary tuberculosis in Ethiopia. The Pan African medical journal 2014;19:162. doi: 10.11604/pamj.2014.19.162.3658 [published Online First: 2014/01/01]

35. Frascella B, Richards AS, Sossen B, et al. Subclinical tuberculosis disease - a review and analysis of prevalence surveys to inform definitions, burden, associations and screening methodology. Clinical Infectious Diseases 2020 doi: 10.1093/cid/ciaa1402

