## Supplementary file S2 for "Tuberculosis prevalence and incidence rates from repeated population-based screening in a district in Ethiopia: a prospective cohort study"

Supplentary file S2 Adult symptom screening questionnaire.

**APPENDIX 2: Adult – tuberculosis symptom based questionnaire – Sidama cluster-study**

**Questionnaire n°.** ☐☐☐☐ **Name of interviewer** ____________________

**Date** ☐☐.☐☐.☐☐ (dd.mm.yy)

---

**1. Socio-demographic variables**

1.1 Name ____________________ 1.2. Date of birth: ☐☐.☐☐.☐☐

1.3. Gender (male=1, female=2) ☐ 1.4.Cellphone. ☐☐☐☐☐☐☐☐☐

1.5. Address: Woreda ____________ Kebele ____________ Cluster ____________

1.6. Marital status: (single=1, married=2, divorced=3, widowed=4, other=9) ☐

If other, specify: ____________________

1.7. Total number of people in the household: ☐☐

Among them, number of children < 5 years of age: ☐☐

Their age(s): ☐☐, ☐☐, ☐☐, ☐☐, ☐☐

1.8. Highest grade completed education: ☐☐,

No schooling (yes=1, no=0. other=9) ☐

If other, specify ____________________

1.9. Occupation:

(farmer=1, housewife= 2, merchant=3, student=4, government employee=5, daily labourer=6, other=9) ☐

If other,(specify) ____________________

**2. Socio-economic variables**

2.1. Number of rooms in the house: ☐☐

2.2. Wall type: (wood with mud/cement or brick=1, wood only=2, other=9) ☐

If other, specify____________________

2.3. Roof type: (corrugated iron sheet=1, thatched/leaf=2, other=9) ☐

If other, specify: ____________________

2.4. Type of fuel for cooking

(electricity=1, kerosone=2, charcoal=3, wood=4, cow doug=5, agriculture by-product=6, no cooking in the household=7, other=9): ☐

If other, specify: ____________________

2.5. Light source: Electricity (yes=1, no=0, other=9): ☐

If other, specify: ______________________________

2.6. Variables for household wealth index (present=1, absent=0, unknown=9)

| Household | | Present | Absent | Unknown |
|---|---|---|---|---|
| 2.6.1 | Separate kitchen in the household | | | |
| 2.6.2 | Cooking room ventilation | | | |
| 2.6.3. | Heating in the house | | | |
| 2.6.4. | Radio | | | |
| 2.6.5. | Television | | | |
| 2.6.6. | Mobile phone | | | |
| 2.6.7. | Refrigerator | | | |
| 2.6.8. | Land for agriculture | | | |
| 2.6.9. | Bank account | | | |

**3. Symptoms of tuberculosis (yes=1, no=0)**

| Symptoms | | Yes | No | Comment (i.e. duration in weeks, dates. enter NK if not known) |
|---|---|---|---|---|
| 3.1. | How long have you been coughing? | / | / | |
| 3.2 | Is this cough productive of sputum? | | | |
| 3.3. | Does the sputum contain blood? | | | |
| 3.4. | Do you have fever? | | | |
| 3.5. | Do you have night sweats? | | | |
| 3.6. | Have you lost your appetite? | | | |
| 3.7. | Have you lost weight? | | | |
| 3.8. | Do you have chest pain or difficulty of breathing? | | | |
| 3.9. | Did you visit a health facility for your current illness? | | | |
| 3.10. | If no in 3.9, reasons for not seeking health care (enter 1 in any applicable):<br><br>Not knowing that it could be TB ☐, not knowing where to go for care ☐, distance to health facility ☐, having to take transport ☐, getting permission to go for care ☐ costs ☐, other ☐<br><br>If other specify: ______________________________ | | | |

**4. Contact-history and risk-factors (yes=1, no=0)**

| Contact-history and risk-factors | | Yes | No | Comment (i.e. duration in weeks, dates. enter NK if not known) |
|---|---|---|---|---|
| 4.1. | Were you treated for tuberculosis before? | | | |
| 4.2. | Any TB case in the household in the past 5 years | | | |
| 4.3. | Did you live with a person who has a chronic cough? | | | |
| 4.4. | Were you tested for HIV in the past year | | | |
| 4.5. | Ever alcohol-drinker | | | |
| 4.6. | Ever chewed Khat | | | |
| 4.7. | Ever smoker | | | |
| 4.8. | Smoker currently in the household | | | |
| 4.9. | Smoker previously in the household | | | |

**5. Clinical and diagnostic information**

5.1. Height (cm) ☐☐☐ 2.2. Weight (kg) ☐☐.☐ 2.3. MUAC (cm): ☐☐

5.2. BCG-scar (yes=1, no=0, unknown=9): ☐

5.3. Sputum sample collection/s

Date of test I: ☐☐.☐☐.☐☐, II: ☐☐.☐☐.☐☐, III: ☐☐.☐☐.☐☐

5.4. Date of smear result: ☐☐.☐☐.☐☐,

Result (positive=1, negative=0): ☐, if positive, grade: ☐☐☐

5.5. Date of GeneXpert result: ☐☐.☐☐.☐☐,

5.6. GeneXpert detection of *M. tb complex* (yes=1, no=0): ☐

5.7. GeneXpert detection of rifampicin-resistance (yes=1, no=0): ☐

5.8. Date of culture result: ☐☐.☐☐.☐☐,

5.9. Detection of *M. tb complex* (yes=1, no=0): ☐

**6. Treatment for tuberculosis disease and follow-up**

6.1. Date of registration: ☐☐.☐☐.☐☐(dd.mm.yy)

6.2. Date of treatment initiation: ☐☐.☐☐.☐☐ (dd.mm.yy)

6.3.Treatment outcome: (cured=1, completed=2, failure=3, death=4, default=5, transfer out=6) ☐, Other, specify:________________________________________

**7. Contact screening**

In household with symptomatic cases, is a contact form with names and age of household contacts filled: Yes ☐ No ☐
