## Supplementary Tables 3 for "Tuberculosis prevalence and incidence rates from repeated population-based screening in a district in Ethiopia: a prospective cohort study"

**Supplementary Tables S3**

Table S1. Description of screening flow with TB patients identified among presumptive TB in Dale 2016-17.

| R1 | Presumptive | All PTB | PTB b+ | PTB s+ | SM- | SM unknown | PTB c+ | EP | No TB |
|---|---|---|---|---|---|---|---|---|---|
| New presumptive only tested once R1 | 1729 | 82 | 71 | 68 | 1658 | 3 | 5 | 6 | 1647 |
| New presumptive with a repeat test R1 | 139 | 46 | 27 | 25 | 113 | 1 | 19 | 0 | 93 |
| Picked up again in R2 or R3 | 184 | 0 | 0 | 0 | 184 | 0 | 0 | 0 | 184 |
| Sum new presumptive R1 | 2052 | 128 | 98 | 93 | 1955 | 4 | 24 | 6 | 1924 |
| R2 | | | | | | | | | |
| New presumptive only tested once R2 | 630 | 75 | 71 | 66 | 560 | 4 | 1 | 3 | 555 |
| New presumptive with a repeat test R2 | 88 | 53 | 19 | 11 | 75 | 2 | 30 | 4 | 35 |
| Prev. presumptive R2 (1, 2) | 166 | 40 | 19 | 15 | 150 | 1 | 19 | 2 | 126 |
| Picked up again in R3 | 35 | 0 | 0 | 0 | 35 | 0 | 0 | 0 | 35 |
| Sum new presumptive R2 | 753 | 128 | 90 | 77 | 670 | 6 | 31 | 7 | 625 |
| Sum prev. presumptive R2 | 166 | 40 | 19 | 15 | 150 | 1 | 19 | 2 | 126 |
| Sum R2 | 919 | 168 | 109 | 92 | 820 | 7 | 50 | 9 | 751 |
| R3 | | | | | | | | | |
| New presumptive only tested once R3 | 900 | 123 | 119 | 106 | 794 | 0 | 2 | 2 | 777 |
| New presumptive with a repeat test R3 | 45 | 14 | 11 | 10 | 35 | 0 | 3 | 0 | 31 |
| Prev. presumptive R3, all | 49 | 26 | 15 | 14 | 35 | 0 | 11 | 0 | 23 |
| R1, R3 | 13 | 6 | 5 | 5 | 8 | 0 | 1 | 0 | 7 |
| R2, R3 | 31 | 18 | 9 | 8 | 23 | 0 | 9 | 0 | 13 |
| R1, R2, R3 | 5 | 2 | 1 | 1 | 4 | 0 | 1 | 0 | 3 |
| Sum new presumptive R3 | 945 | 137 | 130 | 116 | 829 | 0 | 5 | 2 | 808 |
| Sum prev. presumptive R3 | 49 | 26 | 15 | 14 | 35 | 0 | 11 | 0 | 23 |
| Sum R3 | 994 | 163 | 145 | 130 | 864 | 0 | 16 | 2 | 831 |
| Overall | | | | | | | | | |
| New presumptive | 3750 | 393 | 318 | 286 | 3454 | 10 | 60 | 15 | 3357 |
| Pre. presumptive | 215 | 66 | 34 | 29 | 185 | 1 | 30 | 2 | 149 |
| Sum study period, presumptive episodes | 3965 | 459 | 352 | 315 | 3639 | 11 | 90 | 17 | 3506 |
| Sum presumptive episodes | 3965 | | | | | | | | |

*R, round; PTB, PTB b+, bacteriologically diagnosed pulmonary TB; PTB c+, clinically diagnosed pulmonary TB; EP, extra pulmonary TB, PTB s+, smear positive; SM-,smear negative, SM, smear*

Table S2. Screening prevalence of TB in round 1 by catchment area in Dale from October 2016-Jan 2017.

| Catchments | ≥15 age population | Presumptive TB | | PTB s+ | | PTB b+ | | PTB | |
|---|---|---|---|---|---|---|---|---|---|
| | | n | Per 100,000 | n | Per 100,000 | n | Per 100,000 | n | Per 100,000 |
| Mesenkala | 15013 | 238 | 1585(1394-1794) | 10 | 67(34-119) | 10 | 67(34-119) | 15 | 100(58-161) |
| Magara | 10772 | 117 | 1086(902-1298) | 11 | 102(54-177) | 11 | 102(54-177) | 14 | 130(74-212) |
| Hida | 5899 | 63 | 1067(828-1355) | 4 | 68(21-163) | 5 | 85(31-188) | 6 | 102(41-211) |
| Bera | 15855 | 218 | 1375(1202-1565) | 8 | 50(23-96) | 9 | 57(28-104) | 13 | 82(46-136) |
| Goida | 11808 | 151 | 1279(1087-1494) | 6 | 51(20-105) | 6 | 51(20-105) | 8 | 68(31-128) |
| Boa | 18233 | 362 | 1985(1790-2196) | 8 | 44(20-83) | 9 | 49(24-91) | 14 | 77(43-126) |
| Dagiya | 14840 | 222 | 1496(1310-1701) | 12 | 81(44-138) | 12 | 81(44-138) | 12 | 81(44-138) |
| Gidamo | 7433 | 161 | 2166(1853-2516) | 13 | 175(97-291) | 14 | 188(107-308) | 14 | 188(107-308) |
| Moto | 24732 | 300 | 1213(1082-1355) | 13 | 52(29-87) | 13 | 52(29-87) | 14 | 57(32-93) |
| kege | 11598 | 220 | 1897(1660-2158) | 8 | 69(32-131) | 9 | 77(38-142) | 12 | 103(56-175) |
| Total | 136181 | 2052 | 1507(1442-1572) | 93 | 68(54-82) | 98 | 72(58-86) | 122 | 89(74-105) |

*PTB s+, smear positive; PTB b+, bacteriologically diagnosed pulmonary TB; PTB, pulmonary TB*

Table S3. Proportion of pulmonary tuberculosis detected by screening and already on TB treatment at screening, by *Kebele* Dale 2016-2017.

| Catchment HFs | PTB s+ | | | PTB b+ | | | PTB c+ | | | total detected | % detected by screening | | | |
|---|---|---|---|---|---|---|---|---|---|---|---|---|---|---|
| | before | during | total | before | during | total | before | during | total | | PTB s+ | PTB b+ | PTB c+ | total |
| Mesenkala | 17 | 10 | 27 | 17 | 10 | 27 | 7 | 5 | 12 | 39 | 37% (18-66) | 37% (18-66) | 42% (15-92) | 38% (22-62) |
| Megara | 4 | 11 | 15 | 4 | 11 | 15 | 5 | 6 | 11 | 26 | 73% (38-127) | 73% (38-127 | 54% (22-113) | 65% (39-102) |
| Hida | 2 | 4 | 6 | 2 | 5 | 7 | 0 | 0 | 0 | 7 | 67% (21-160) | 71% (26-158) | - | 71% (26-158) |
| Moto | 3 | 8 | 11 | 3 | 9 | 12 | 2 | 2 | 4 | 16 | 73% (33-138) | 75% (16-83) | 50% (8-165) | 69% (36-119) |
| Dagiya | 9 | 6 | 15 | 9 | 6 | 15 | 0 | 0 | 0 | 15 | 40% (16-83) | 40% (16-83) | - | 40% (16-83) |
| Gidamo | 5 | 8 | 13 | 5 | 9 | 14 | 0 | 0 | 0 | 14 | 61% (28-116) | 64% (31-118) | - | 64% (31-118) |
| Goida | 3 | 12 | 15 | 3 | 12 | 15 | 8 | 4 | 12 | 27 | 80% (43-136) | 80% (43-136) | 33% (10-80) | 59% (35-94) |
| Bera | 4 | 13 | 17 | 4 | 14 | 18 | 6 | 4 | 10 | 28 | 76% (42-127) | 78% (44-127) | 40% (13-96) | 64% (39-100) |
| Boa | 2 | 13 | 15 | 2 | 13 | 15 | 1 | 2 | 3 | 18 | 87% (48-144) | 87% (48-144) | 67% (11-220) | 83% (48-134) |
| kege | 0 | 8 | 8 | 0 | 9 | 9 | 1 | 1 | 2 | 11 | 100% (46-190) | 100% (46-190) | 50% (2.5-246) | 91% (46-162) |
| Grand total | 49 | 93 | 142 | 49 | 98 | 147 | 30 | 24 | 54 | 201 | 65% (53-80) | 67% (54-80) | 44% (29-65) | 60% (50-72) |

*HFs, health facilities; PTB s+, smear-positive; PTB b+, bacteriologically diagnosed pulmonary TB (including PTB s+); PTB c+, clinically diagnosed pulmonary TB*

Table S4. Proportion of pulmonary tuberculosis detected by screening and already on TB treatment at screening, by age and sex Dale 2016-2017.

| Covariates | PTB among presumptive TB identified during screening | | | | Proportion presumptive with PTB | | | PTB already on anti-TB in round at screening | | | | % Detected by screening | | | |
|---|---|---|---|---|---|---|---|---|---|---|---|---|---|---|---|
| | Presumptive TB | PTB s+ | PTB b+ | PTB c+ | PTB s+ | PTB b+ | PTB c+ | PTB s+ | PTB b+ | PTB c+ | Total detected | PTB s+ | PTB b+ | PTB c+ | total |
| Total | 2052 | 93 | 98 | 24 | 5% | 5% | 1% | 49 | 49 | 30 | 201 | 65% | 67% | 44% | 61% |
| Age | | | | | | | | | | | | | | | |
| 15-24 | 294 | 25 | 27 | 7 | 9% | 9% | 2% | 23 | 23 | 5 | 62 | 52% | 54% | 58% | 55% |
| 25-34 | 395 | 30 | 31 | 7 | 8% | 8% | 2% | 15 | 15 | 6 | 59 | 67% | 67% | 54% | 64% |
| 35-44 | 437 | 26 | 26 | 2 | 6% | 6% | 0% | 4 | 4 | 2 | 34 | 87% | 87% | 50% | 82% |
| 45-54 | 432 | 5 | 7 | 4 | 1% | 2% | 1% | 3 | 4 | 5 | 20 | 62% | 64% | 44% | 55% |
| 55+ | 494 | 7 | 7 | 4 | 1% | 1% | 1% | 4 | 3 | 12 | 26 | 63% | 70% | 25% | 42% |
| Sex | | | | | | | | | | | | | | | |
| Male | 847 | 52 | 56 | 15 | 6% | 7% | 2% | 27 | 27 | 17 | 115 | 66% | 67% | 47% | 62% |
| Female | 1205 | 41 | 42 | 9 | 3% | 3% | 1% | 22 | 22 | 13 | 86 | 65% | 66% | 41% | 59% |

*PTB s+, smear-positive; PTB b+, bacteriologically diagnosed pulmonary TB (including PTB s+); PTB c+, clinically diagnosed pulmonary TB*

Table S5. Screening and overall prevalence of pulmonary TB (including PTB s+) in round 1, by age and sex in Dale 2016-17.

| Covariates | Population | Prevalence of PTB identified during screening | | | | | | Overall prevalence of PTB (including already those on treatment) | | | | | |
|---|---|---|---|---|---|---|---|---|---|---|---|---|---|
| | | PTB s+ | per 100,000 | PTB b+ | per 100,000 | PTB c+ | per 100,000 | PTB s+ | per 100,000 | PTB b+ | per 100,000 | PTB c+ | per 100,000 |
| Total | 136181 | 93 | 68(54-82) | 98 | 72(57-86) | 24 | 17(10-24) | 142 | 104(87-121) | 147 | 108(90-125) | 54 | 39(29-50) |
| Age | | | | | | | | | | | | | |
| 15-24 | 49735 | 25 | 50(33-73) | 27 | 54(37-77) | 7 | 17(9-27) | 48 | 96(72-127) | 50 | 100(76-132) | 12 | 24(13-41) |
| 25-34 | 33958 | 30 | 88(60-125) | 31 | 91(63-128) | 7 | 21(9-41) | 45 | 132(97-176) | 46 | 135(100-179) | 13 | 38(21-64) |
| 35-44 | 23797 | 26 | 109(73-158) | 26 | 109(73-158) | 2 | 8(1.3-28) | 30 | 126(86-177) | 30 | 126(86-177) | 4 | 17(5-40) |
| 45-54 | 13369 | 5 | 37(14-83) | 7 | 52(22-104) | 4 | 21(8-41) | 8 | 60(27-114) | 11 | 82(43-143) | 9 | 67(33-123) |
| 55+ | 15319 | 7 | 46(20-90) | 7 | 45(20-90) | 4 | 26(9-63) | 11 | 72(38-125) | 10 | 65(33-116) | 16 | 104(61-166) |
| Sex | | | | | | | | | | | | | |
| Male | 67681 | 52 | 77(58-100) | 56 | 83(63-107) | 15 | 22(12-35) | 79 | 117(93-144) | 83 | 122(98-151) | 32 | 47(33-66) |
| Female | 68500 | 41 | 60(43-80) | 42 | 61(44-82) | 9 | 13(6-24) | 63 | 92(71-117) | 64 | 93(72-118) | 22 | 32(21-48) |

*PTB s+, smear-positive; PTB b+, bacteriologically diagnosed pulmonary TB (including PTB s+); PTB c+, clinically diagnosed pulmonary TB*

Table S6. Incidence and proportion of smear-positive TB in the adult population in Round 2+3, by age and sex Dale, Feb-Sep 2017.

| Covariates | Person-years | *Presumptive TB, n* | PTB s+, *n* | Incidence per 100,000 | Proportion of presumptive TB with PTB s+ *(%)* |
|---|---|---|---|---|---|
| Overall | 96388 | 1909 | 222 | 230(201-262) | 12% |
| Age in years | | | | | |
| 15-24 | 35063 | 338 | 78 | 222(177-276) | 23% |
| 25-34 | 23973 | 465 | 85 | 354(285-436) | 18% |
| 35-44 | 16879 | 362 | 29 | 171(117-244) | 8% |
| 45-54 | 10918 | 367 | 14 | 128(73-210) | 4% |
| 55+ | 9555 | 377 | 16 | 167(99-266) | 4% |
| Sex | | | | | |
| Male | 47823 | 826 | 108 | 226(187-271) | 13% |
| Female | 48565 | 1083 | 114 | 235(195-281) | 11% |

*PTB s+, smear-positive; PTB b+; TB, tuberculosis*

Table S7. Case Notification per 100,000 of bacteriologically diagnosed pulmonary TB in Dale 2015/16-2017/18.

| HFS | Before-screening | | | | | | By screening | | | | | | After screening | | | | | |
|---|---|---|---|---|---|---|---|---|---|---|---|---|---|---|---|---|---|---|
| | 2015_ q4 | 2016 _q1 | 2016_ q2 | 2016 _q3 | Total | per 100000 | 2016_ q4 | 2017 _q1 | 2017_ q2 | 2017_ q3 | Tota l | per 100,000 | 2017_ q4 | 2018 _q1 | 2018_ q2 | 2018 _q3 | Tot al | per 100,000 |
| Mesenkala | 4 | 5 | 10 | 7 | 26 | 174 | 10 | 8 | 11 | 8 | 37 | 246 | 6 | 2 | 4 | 5 | 17 | 108 |
| Megara | 4 | 5 | 3 | 1 | 13 | 121 | 14 | 6 | 5 | 5 | 30 | 278 | 3 | 6 | 7 | 5 | 21 | 185 |
| Hida | 0 | 0 | 0 | 2 | | 0 | 6 | 2 | 4 | 4 | 16 | 271 | 0 | 0 | 0 | 0 | 0 | 0 |
| Moto | 1 | 1 | 2 | 1 | 5 | 20 | 16 | 5 | 5 | 3 | 29 | 117 | 6 | 11 | 10 | 10 | 37 | 142 |
| Dagiya | 2 | 6 | 4 | 5 | 17 | 115 | 13 | 11 | 5 | 5 | 34 | 229 | 3 | 12 | 4 | 7 | 26 | 167 |
| Gidamo | 5 | 3 | 2 | 3 | 13 | 175 | 8 | 14 | 3 | 13 | 38 | 514 | 4 | 4 | 6 | 7 | 21 | 271 |
| Goida | 4 | 0 | 0 | 3 | 7 | 59 | 7 | 7 | 5 | 14 | 34 | 288 | 8 | 5 | 4 | 7 | 24 | 193 |
| Bera | 2 | 6 | 2 | 2 | 12 | 76 | 8 | 13 | 10 | 16 | 47 | 296 | 6 | 4 | 4 | 3 | 17 | 102 |
| Boa | 0 | 8 | 0 | 2 | 10 | 55 | 8 | 15 | 9 | 22 | 54 | 297 | 10 | 7 | 14 | 3 | 34 | 178 |
| kege | 2 | 3 | 0 | 0 | 5 | 43 | 7 | 13 | 7 | 6 | 33 | 286 | 3 | 4 | 6 | 9 | 22 | 182 |
| Gran total | 24 | 37 | 23 | 26 | 110 | 81 | 98 | 94 | 64 | 96 | 352 | 258 | 49 | 55 | 59 | 56 | 219 | 153 |

Table S8. Case notification per 100,000 of clinically diagnosed pulmonary TB in Dale 2015/16-2017/18.

| HFS | Before-screening | | | | | | By screening | | | | | | After screening | | | | | |
|---|---|---|---|---|---|---|---|---|---|---|---|---|---|---|---|---|---|---|
| | 2015_ q4 | 2016 _q1 | 2016 _q2 | 2016_ q3 | Total | per 100000 | 2016 _q4 | 2017_ q1 | 2017 _q2 | 2017 _q3 | Total | per 100,000 | 2017 _q4 | 2018_ q1 | 2018 _q2 | 2018 _q3 | Total | per 100,000 |
| Mesenkala | 5 | 6 | 1 | 7 | 19 | 127 | 5 | 4 | 7 | 0 | 16 | 106 | 0 | 1 | 4 | 2 | 7 | 44 |
| Megara | 4 | 2 | 3 | 4 | 13 | 121 | 6 | 4 | 2 | 1 | 13 | 121 | 1 | 0 | 0 | 0 | 1 | 9 |
| Hida | 0 | 0 | 0 | 0 | 0 | 0 | 0 | 0 | 0 | 0 | 0 | | 0 | 0 | 0 | 0 | 0 | 0 |
| Moto | 0 | 0 | 0 | 2 | 2 | 8 | 2 | 5 | 17 | 1 | 25 | 101 | 3 | 0 | 4 | 0 | 7 | 27 |
| Dagiya | 0 | 0 | 0 | 0 | 0 | 0 | 0 | 0 | 0 | 0 | 0 | | 0 | 0 | 0 | 0 | 0 | 0 |
| Gidamo | 2 | 0 | 0 | 0 | 2 | 27 | 0 | 0 | 0 | 0 | 0 | | 0 | 0 | 0 | 0 | 0 | 0 |
| Goida | 1 | 5 | 5 | 5 | 16 | 136 | 4 | 0 | 0 | 0 | 4 | 34 | 0 | 0 | 1 | 1 | 2 | 16 |
| Bera | 8 | 3 | 3 | 3 | 17 | 108 | 4 | 4 | 2 | 11 | 21 | 132 | 0 | 1 | 0 | 1 | 2 | 12 |
| Boa | 0 | 0 | 1 | 0 | 1 | 6 | 2 | 1 | 0 | 0 | 3 | 16 | 0 | 0 | 0 | 0 | 0 | 0 |
| kege | 0 | 0 | 0 | 1 | 1 | 9 | 1 | 2 | 0 | 5 | 8 | 69 | 1 | 1 | 1 | 0 | 3 | 25 |
| Gran total | 20 | 16 | 13 | 22 | 71 | 52 | 24 | 20 | 28 | 18 | 90 | 66 | 5 | 3 | 10 | 4 | 22 | 15 |

Table S9. Case notification per 100,000 of smear-positive pulmonary TB in Dale 2015/16-2017/18.

| HFS | Before-screening | | | | | | Screening | | | | | | After screening | | | | | |
|---|---|---|---|---|---|---|---|---|---|---|---|---|---|---|---|---|---|---|
| | 2015_ q4 | 2016_ q1 | 2016 _q2 | 201 6_q3 | Total | per 10000 0 | 2016_ q4 | 2017_ q1 | 2017_ q2 | 2017 _q3 | Total | per 100,000 | 2017_ q4 | 2018 _q1 | 2018_ q2 | 2018 _q3 | Total | per 100,000 |
| Mesenkala | 4 | 5 | 10 | 7 | 26 | 174 | 12 | 8 | 8 | 7 | 35 | 233 | 6 | 2 | 4 | 5 | 17 | 108 |
| Megara | 4 | 5 | 3 | 1 | 13 | 121 | 8 | 9 | 5 | 4 | 26 | 241 | 3 | 6 | 7 | 5 | 21 | 185 |
| Hida | 0 | 0 | 0 | 2 | | 0 | 5 | | 4 | 4 | 13 | 220 | 0 | 0 | 0 | 0 | 0 | 0 |
| Moto | 1 | 1 | 2 | 1 | 5 | 20 | 12 | 5 | 5 | 3 | 25 | 101 | 6 | 11 | 9 | 10 | 36 | 138 |
| Dagiya | 2 | 6 | 4 | 5 | 17 | 115 | 11 | 13 | 5 | 5 | 34 | 229 | 3 | 11 | 4 | 7 | 25 | 160 |
| Gidamo | 5 | 3 | 2 | 3 | 13 | 175 | 12 | 9 | 3 | 8 | 32 | 433 | 4 | 4 | 6 | 7 | 21 | 271 |
| Goida | 4 | 0 | 0 | 3 | 7 | 59 | 7 | 11 | 5 | 10 | 33 | 279 | 8 | 5 | 4 | 7 | 24 | 193 |
| Bera | 2 | 6 | 2 | 2 | 12 | 76 | 11 | 9 | 10 | 10 | 40 | 252 | 6 | 4 | 4 | 3 | 17 | 102 |
| Boa | 0 | 8 | 0 | 2 | 10 | 55 | 8 | 9 | 9 | 20 | 46 | 253 | 9 | 6 | 14 | 3 | 32 | 167 |
| kege | 2 | 3 | 0 | 0 | 5 | 43 | 7 | 13 | 6 | 5 | 31 | 269 | 3 | 4 | 6 | 9 | 22 | 182 |
| Gran total | 24 | 37 | 23 | 26 | 110 | 81 | 93 | 86 | 60 | 76 | 315 | 231 | 48 | 53 | 58 | 56 | 215 | 150 |
